# Tailored Food is Medicine Programs as an effective approach to address dietary intake and blood pressure among rural and urban adults

**DOI:** 10.1101/2025.07.09.25331229

**Authors:** Alison Gustafson, Carolyn Lauckner, Joshua Bush, Christa Mayfield, Emily Dimond, Carolina Morales, Elizabeth T. Anderson Steeves

## Abstract

**Background:** Food is Medicine (FIM) programs have shown potential at improving health outcomes, reducing food insecurity, and increasing dietary intake. However, few interventions have sought to individually tailor these programs based on user preferences and constraints. This program utilized a screening decision tool to allocate adults to a tailored FIM program to examine process and clinical outcomes.

**Methods:** Adults ages 18-64 with hypertension were screened for food insecurity at two large hospital systems (one rural, one urban) in Kentucky. Participants who screened positively and wanted assistance with food were referred to the Food as Health Alliance hub to receive either medically tailored meals or grocery prescription (Rx). Medically tailored meals (MTM) provided 5 meals per week for 12 weeks. The grocery Rx program provided $100 each month for 3 months to purchase food consistent with guidelines for people with hypertension. Baseline and post-intervention outcomes were obtained from electronic medical records, and process measures included engagement, dose, and program acceptability. Semi-structured interviews were completed with a subset of 20 participants to obtain qualitative feedback on the program.

**Results:** A total of 159 participants referred were enrolled, and 144 participants completed all measures (complete case rate of 91%). There were no significant changes within grocery Rx or MTM for the primary outcome of systolic or diastolic blood pressure from baseline to post intervention. However, there were significant effects on dietary intake, financial strain, and self-reported general health among grocery Rx and MTM recipients. Among those receiving Supplemental Nutrition Assistance Program (SNAP) benefits who received grocery Rx, there was a significant change in systolic (-8.75 95% CI-16.83,-.67) and diastolic blood pressure (-5.42 95% CI-10.72,-.13) relative to those not receiving SNAP. Qualitative findings aligned with the quantitative findings in that participants reported high levels of satisfaction and that the program allowed them to eat healthier, helped ease the burdensome cost of food, and improved various aspects of their health.

**Conclusion:** A tailored FIM program can improve dietary intake and reduce blood pressure among key subpopulations participating in the program in the short-term.

Clinical trial registration NCT07011251

## Introduction

Across the United States, food insecurity rates have doubled between 2005 and 2021^1^. In addition, millions more report experiencing nutrition insecurity, defined as lack of consistent affordable access to nutritious foods and beverages that promote health^2^.

Food and nutritional insecurity are associated with increased risk of cardiovascular disease^3^, hypertension, and diabetes^4^. Those from lower socio-economic statuses experience higher rates of food insecurity and diet-sensitive chronic disease^5–9^.

Currently, several federal food programs have aimed to address food insecurity through providing supplemental funds to help families or individuals receive additional food, such as Farmers’ Market Nutrition Program or Congregate Meals Program. The largest federal safety net program is the Supplemental Nutrition Assistance Program (SNAP), which provides on average $183 per month per qualifying individual for food-related purchases^10^. While there is a body of research indicating SNAP has been successful at addressing food insecurity,^11–13^ SNAP is not explicitly intended to encourage healthy food options that are tailored for specific diet-sensitive chronic disease. Thus, the Food is Medicine (FIM) movement was created to address the complexity of food and nutrition security among food insecure and lower income households^14^.

Many states have launched FIM programs through an 1115 Medicaid Waiver or through In Lieu of Service protocols^15^. These food and nutrition services provide medically tailored meals (MTM), grocery prescriptions (Rx), meal boxes, or produce prescriptions for those enrolled in Medicaid and who meet other eligibility criteria. These programs are currently offered in select states and among the insured. Preliminary studies have found that FIM has reduced food insecurity, improved health outcomes, and dietary quality^16–18^. Meanwhile, others have found no significant change in health outcomes^19^. In general, results thus far have found a reduction in healthcare costs, fewer hospital admissions, and fewer admissions to skilled nursing^14,20^. A key limitation of some of this work is the lack of tailored programs to meet patient needs and preferences (e.g., cooking ability, access to a full kitchen), which might allow greater engagement and improved outcomes. In some FIM programs, activation of subsidy cards has been below 50%^21,22^, and in healthy food incentive programs, like the US Department of Agriculture Gus Schumacher Nutrition Incentive Program (GusNIP) produce prescription program, only about 65% of the card balances are used^23^. These low engagement rates may indicate that there is no one-size-fits-all approach and that offering only one type of program that is not tailored to individuals’ needs will not produce the intended outcomes^24^.

Recently, the American Heart Association (AHA) launched the Health Care by Food™ (HCxF) program, which is advocating for a user-centered design approach. Our study utilized this framework to develop a tailored FIM program using a screening decision tool to allocate patients to a FIM program based on their preferences and resource constraints, while also balancing the cost of these programs. The aims of this study are to: 1) Examine the use of a screening decision tool on process measures of engagement, retention, and usage; 2) Determine effects of the FIM program on primary outcomes of blood pressure (systolic and diastolic) and on secondary outcomes of dietary intake, financial strain, and general self-reported health; 3) Report on cost benefits of the program among participants; 4) Report on the user experience among participants through collection of qualitative feedback, and 5) Assess cost-effectiveness of the program.

### Clinical Perspective

- What is new? The study provides new information related to how to tailor a FIM program based on user needs and preferences for improved health outcomes.
- What are the clinical implications? Practitioners can be a vital partner in screening and referring for FIM programs to help ensure improved health outcomes among patients.

## Methods

The data that support the findings of this study are available from the corresponding author upon reasonable request.

We have described recruitment, enrollment, and eligibility in a previous publication in more detail (Anderson Steeves, PhD, MPH, accepted for publication, JHPCU 2025). Briefly described below are our screening, referral, and enrollment steps.

### Eligibility Criteria

To be eligible to participate in this pilot study, patients had to be between the ages of 18-64, have a diagnosis of hypertension (ICD-10 code I10), and speak conversational English. Only one patient per household was eligible to participate. Participants were recruited over a 45-day period from two large hospital systems in rural and urban communities in Kentucky.

### Screening

Two screening processes, automated or face-to-face clinician-administered, were used based on the current structure and preferences of the health system partners. The automated procedure consisted of the patient receiving an e-mail with instructions to log into their patient portal before check-in and complete the Hunger Vital Signs screener,^25^ with responses recorded in the electronic medical record (EMR). The face-to-face procedure consisted of the nurse or clinic staff asking the patient the Hunger Vital Signs screener in the patient room and documenting their response in the EMR.

### Referral

If the eligible patient expressed interest in participating, the healthcare clinic staff would input their relevant contact and eligibility information into a Research Electronic Data Capture REDCap form^26^ for the Food as Health Alliance team to contact the patient for further enrollment procedures.

### Enrollment into the HCxF program

Staff from the Food as Health Alliance Hub at the University of Kentucky coordinated the enrollment of patients and subsequent delivery of the food package. Referred patients received a welcome message via text and email with a link to the baseline survey and phone number to enroll. Follow-up messages were sent two days later to patients not enrolled, after which staff made three attempts by phone to assist with enrollment. Enrollment consisted of informed consent for the pilot study, use of a screening decision tool to match participants to the appropriate program, and a baseline survey. This study was approved by the University of Kentucky institutional review board (Protocol #93234). All participants completed informed consent through REDCap.

### Development of the Screening Decision Tool

The study team constructed a screening tool that took into account user preferences, such as a desire to shop for food independently or prepare meals, while balancing resource constraints such as having reliable transportation to get to a grocery store. The development of the tool utilized questions from other social determinants of health items.^27,28^ The team pilot tested the screening tool among previous FIM patients in rural and urban areas to understand if the tool was sensitive to preferences and constraints. A total of N=31 pilot tested the tool, which resulted in the following revisions: 1) removed stigmatizing language, such as ‘prepackaged meals;’ 2) asked if participants are *easily* able get to the grocery store rather than just “able.” As one participant noted, she could “*walk to the store on good days, but not always”*; 3) asked participants if they “preferred delivery more than in-sperson shopping” versus “preferred choosing food on their own in store more than delivery.” The original phrasing seemed to be biased toward in-person shopping. The study team made final edits and tested the revised version among N=8 additional participants (see Supplemental Materials for the final screening decision tool) to ensure the tool was functioning as intended and that no additional issues were reported.

## Intervention Content

### Medically Tailored Meals (MTM)

Mom’s Meals was the provider of MTM, which consisted of 5 meals per week for 12 weeks tailored to those with hypertension. The meal options were low in sodium, saturated fat, and met guidelines from AHA. Ten meals were delivered to participants’ homes every other week (6 deliveries) for those that were allocated to this program. Participants could select meals from a menu of options that met dietary guidelines before each delivery.

### Grocery Rx (urban areas)

Kroger, with a partnership from Soda Health for the grocery prescription reloadable cards, was the provider for this arm and consisted of $100 per month for 3 months. The food options were low in sodium and saturated fat and emphasized fruits, vegetables, and low-fat dairy. These options meet criteria from AHA dietary guidelines and were created by Registered Dietitians from the study team and from Kroger Health. The card was sent to participants’ homes at the start of the program, and $100 was reloaded onto their card for the remaining months.

### Grocery Rx (rural areas)

Food City was the provider for this arm and consisted of $100 per month for 3 months in the form of physical vouchers mailed to participants’ homes. The food options were the same as described for the grocery Rx (urban). Four vouchers worth $25 each were mailed to participants each month. Participants were encouraged to use the full amount of funds available from the vouchers when shopping as change could not be given for an unused portion of a single voucher.

### Grocery Rx (online)

Instacart Fresh Funds program was the provider for this arm and has been described previously^15^. This online program offers participants a code to be redeemed online to purchase healthy food items that are delivered to their door.

Participants needed to have a bank account and funds available in that account in order to use this program. The decision tool took into account this user constraint when allocating patients to various programs and thus only one participant had the capacity to utilize this program. Due to Instacart not being available in some rural areas, grocery Rx online was only available to urban participants for this study.

## Data Collection

Clinic nurses uploaded protected health information (PHI) into our referral portal using REDCap, which is HIPAA authorized for PHI. PHI was uploaded at baseline and post intervention including systolic and diastolic blood pressure (primary outcome), height, weight, and hypertension medication use. Patient health data was captured from clinic staff as part of standard of care. Baseline data was defined as the systolic and diastolic captured in the past year. Post intervention data was captured within 45 days of completing the intervention. The study team reminded participants to return to their primary care provider for a follow-up visit to capture PHI at timepoint two.

Patients also completed a baseline and post-intervention survey, which asked questions on all secondary outcomes and process measures. In accordance with the AHA HCxF program, we utilized the common core and preferred measures^29^. Under optional measures, we also collected data on employment and engagement for comparison to other studies. Table 1. provides details regarding timing and exact measures used.

**Table 1.** Outcomes and Process Measures, Instrument, and Timing of Data Collection.

|  |  |  |
| --- | --- | --- |
| **Primary and Secondary Outcome Measure** | **Instrument/Data Source** | **Timing** |
| Biometrics (Blood Pressure, Primary Outcome) | Patient electronic medical record | Baseline, 3-month |
| Food Insecurity<sup>30</sup> | USDA 6-item food security screener | Baseline, 3-month |
| Nutrition Security | 1-item Center for Nutrition and Health Impact | Baseline,3-month |
| Diet Quality | DSQ-10 | Baseline,3-month |
| General Health Status<sup>29</sup> | Single Item: *Would you say that in general your health is excellent, very good, good, fair, or poor?* | Baseline, 3-month |
| Mobility, Self-care, Usual activities, Pain, Depression<sup>29</sup> |  | Baseline, 3-month |
| Financial Strain | Single Item: I don't have money to pay my bills | Baseline, 3-month |
| **Process Measures** |  |  |
| Net Promoter Score | Single Item "How likely is it that you would recommend the [intervention] program to a friend?" | Post Intervention |
| Budget impact | Single Item "How did the program affect your budget?" | Post intervention |
| Engagement (self-report) | Single Item "If you had to pay for this program, would you choose to participate?" | Post Intervention |
| Communication | E-mail, text, and phone calls | 3-months |
| Redemption Rate Grocery Rx | Amount of grocery Rx funds used/amount of funds issued | 3-months |
| Dose MTM | % of meals delivered | 3-months |
| Engagement rate (Objective) | # of enrolled/# that completed the program | 3-months |
| Complete case | # completed program with all measures collected/#enrolled | 3-months |

Everything in Table 1. is a core or preferred measure by AHA.

Semi-structured qualitative interviews were conducted to obtain post-program participant/user perspectives. An interview guide was developed and reviewed by the study team to obtain information related to participant satisfaction with the HCxF program they were assigned to, barriers and facilitators to participating, perceived impacts of the program, and any recommended changes. Interviews were conducted by trained research staff over the phone and audio recorded with permission. Recordings were transcribed verbatim and reviewed by interviewers for accuracy. A purposive sampling strategy for the interviews was used to obtain feedback from the various HCxF prescriptions (MTM, grocery Rx card, and grocery Rx vouchers). The grocery Rx online option was omitted from sample due to the small size (n=1). If selected, the participant was contacted by the research team and invited to participate in the interview.

Participants received a $50 incentive for completing an interview. Recruitment and interview data collection occurred until saturation was reached, which was defined as identifying no new information during the interview^31^. After saturation was reached, an additional four interviews were conducted to confirm saturation, and then data collection was discontinued.

## Analyses

The PI takes full responsibility for the data management and analyses as presented.

Descriptive statistics with mean, SE, and percentage are reported for demographic variables. T-test and chi-square tests were used to look at changes pre-and post-intervention. Linear regression adjusted for sex, age, medication use, household size, household income, and race/ethnicity to examine the effect of participation in the FIM on primary and secondary outcomes. GLM was used to assess mean changes in dietary intake for men and women baseline to post intervention. Dietary intake is analyzed separately for men and women since they have different dietary needs and national guidelines. Chi square analyses were used to measure changes for categories in general health, financial strain, and food security status.

*Semi-structured Qualitative Interview Analysis*. Thematic analysis^32^ was used to identify themes from semi-structured interviews with participants who had recently completed

the program. A codebook was iteratively developed by the coding team (CM, ED, and EAS). The initial codebook was developed by having coders complete line by line coding of two information-rich transcripts to identify axial codes. The axial codes were then discussed by the coders, and codes were defined. The codebook was then uploaded into Dedoose (version 9.2.12, Los Angeles, CA: SocioCultural Research Consultants, LLC) Qualitative Analysis software. Two interrater reliability (IRR) training tests within Dedoose were completed. Tests consistently found a kappa statistic ≥0.6 indicating substantial agreement between coders. After the IRR tests were run and found to be acceptable, the remaining transcripts were independently coded.

Throughout the coding process, the coding team met weekly to discuss coding questions/discrepancies, emergent themes, and memos created during the coding process. Themes were identified from the coded transcripts and summaries with supportive quotes were created for each theme.

The costs related to the interventions include baseline costs that were incurred regardless of the study arm and those that were specific to each arm. Costs related to the training of clinic staff, screening patients for eligibility, referral and enrollment, as well as development and distribution of patient materials are included in the base costs of the intervention. Participant support and assistance efforts were also recorded as total costs, not associated with a specific arm of the study.

## Results

### Screening Decision Tree Results

The full results from the tool indicated N=34 matched to MTM, N=116 matched to in-person grocery Rx, and N=1 matched to online grocery Rx.

### Study Diagram

Figure 1 provides the study flow from screening, referral, enrollment, and engagement over the entire study period.

**Figure 1.**
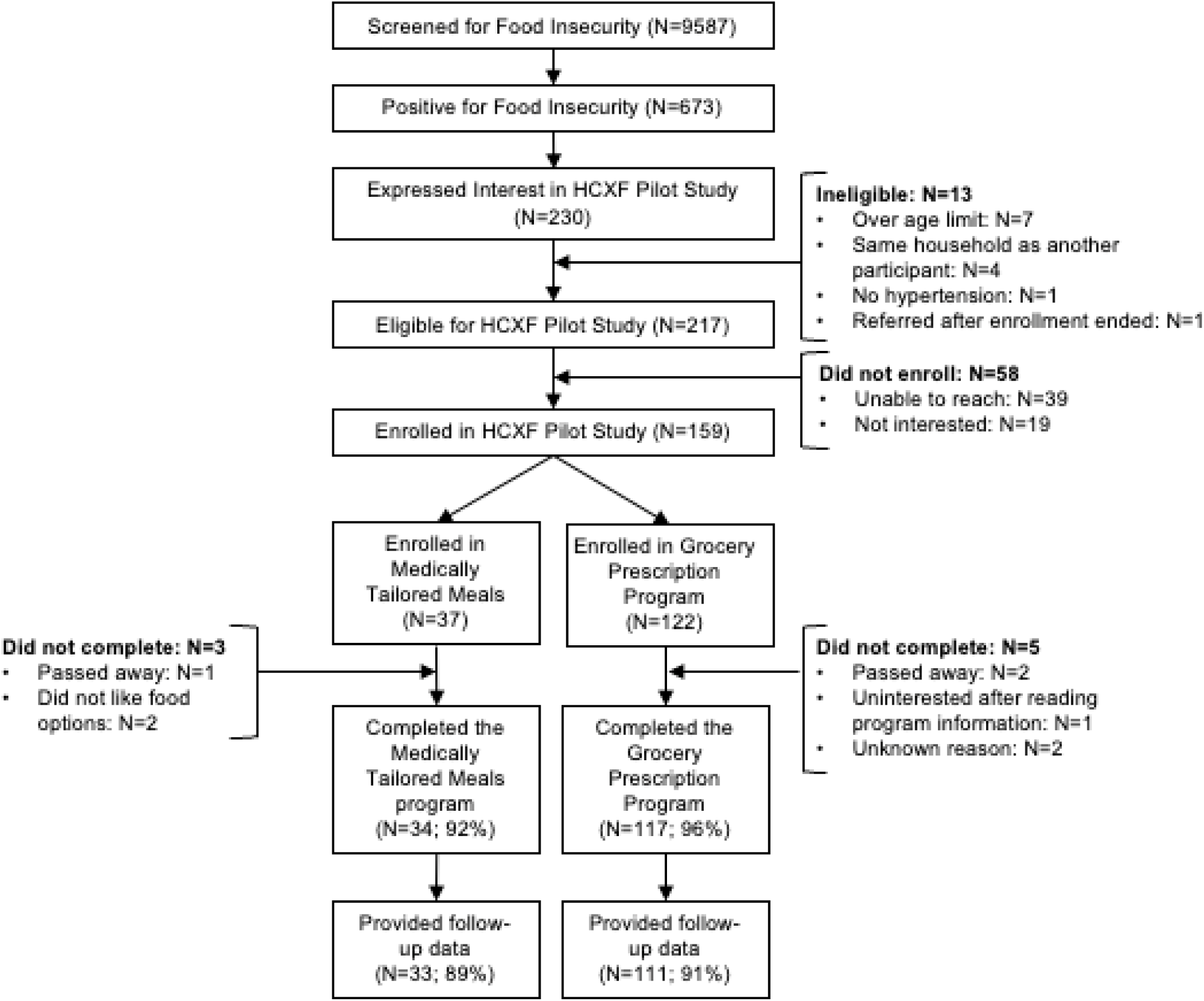
Study flow of the screening, referral, enrollment & engagement process

Of enrolled participants who completed the program, N=7 did not provide health outcomes at post intervention. Of those, 3 were in the rural grocery Rx arm, 3 in the urban grocery Rx, and 1 in the urban MTM.

### Process Measures Results

Table 2. provides key results on process measures across the study period. In general, participants reported a very high net promoter score indicating they are very likely to tell a friend about the program. Close to 40% reported they would pay for this program if it were not free. In addition, our program had a very high redemption rate of close to the full amount of funds being used each month. Almost all those in the MTM completed the program.

**Table 2.** Process Measure Results (N=136 for post survey data collection and N=151 for engagement metrics)

|  |  |
| --- | --- |
| Net Promoter Score (N=136, scale 1-10; not at all likely to very likely) | Mean: 9.45 (95% CI 9.21, 9.69); *Indicates high likelihood of telling a friend about the program* |
| Budget Impact (N=136) | 54% were able to buy additional food<br>54% were able to buy better quality food<br>49% were able to use money typically spent on food for other needs |
| Engagement (Self-report) (N=136) | Yes, would pay: 39%<br>No, would not pay: 37% |
| Communication | On average, participants sent 6 text messages, 1.1 emails, and placed 0.5 phone calls over the study period |
| Redemption Rate (Grocery Rx) (N=151) | Mean: Month 1: \$83; Month 2: \$88, Month 3: \$81<br>Median: Month 1: \$97; Month 2: \$99; Month 3: \$99 |
| Dose MTM (N=151) | 99.5% of intended meals were delivered (Of 34 participants enrolled, 33 received all 6 meal deliveries, 1 received 5 deliveries) |
| Engagement Rate | 151/159 = 95% completed program |
| Complete Case | 144/159 = 91% completed all follow-up measures |

Primary and secondary outcomes are shown in Table 3 and Table 4. The average age of participants was 52, and a majority were White and had at least a High School degree. There was a high unemployment rate of about 78%, and 50% participation in SNAP.

**Table 3.**
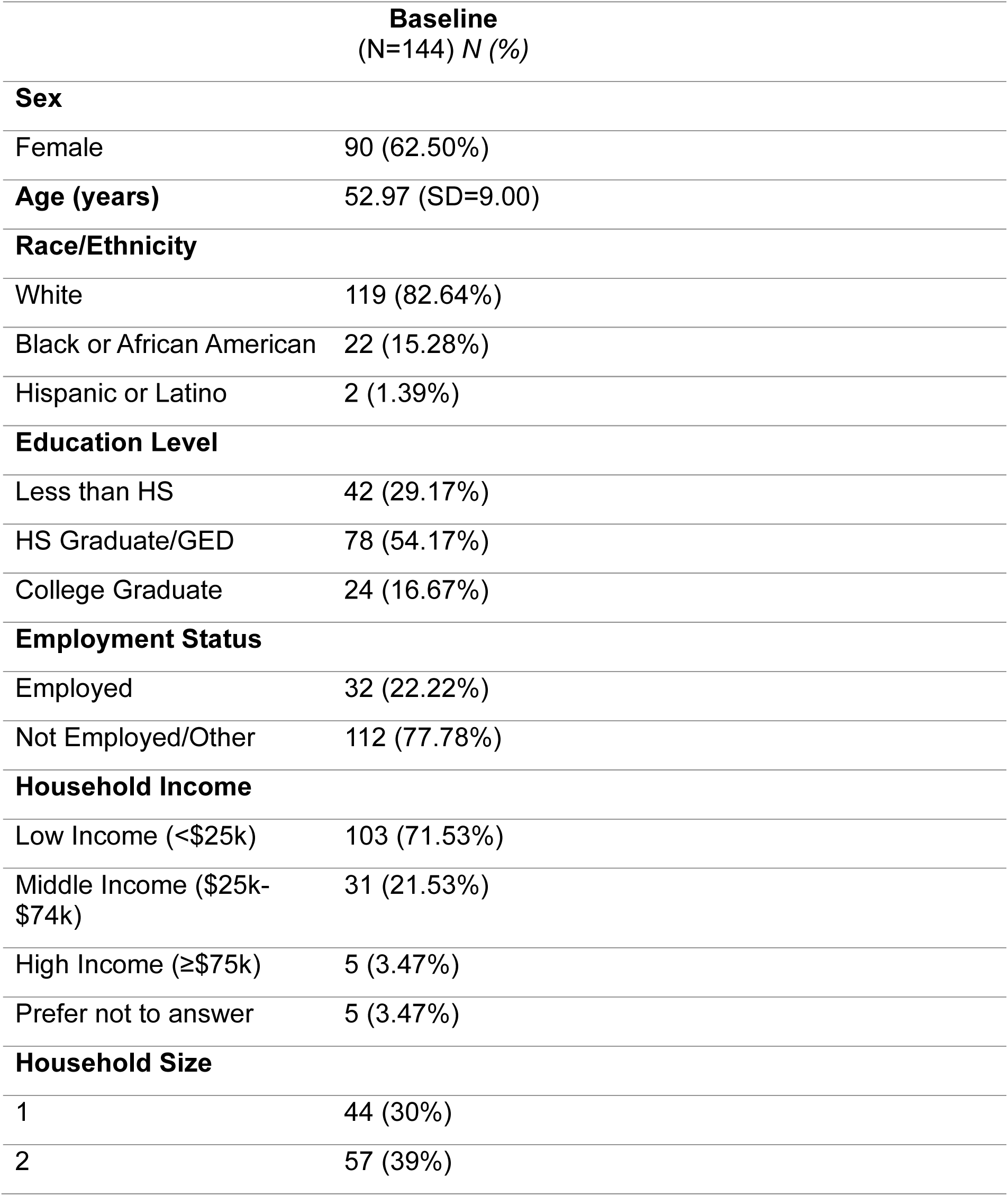
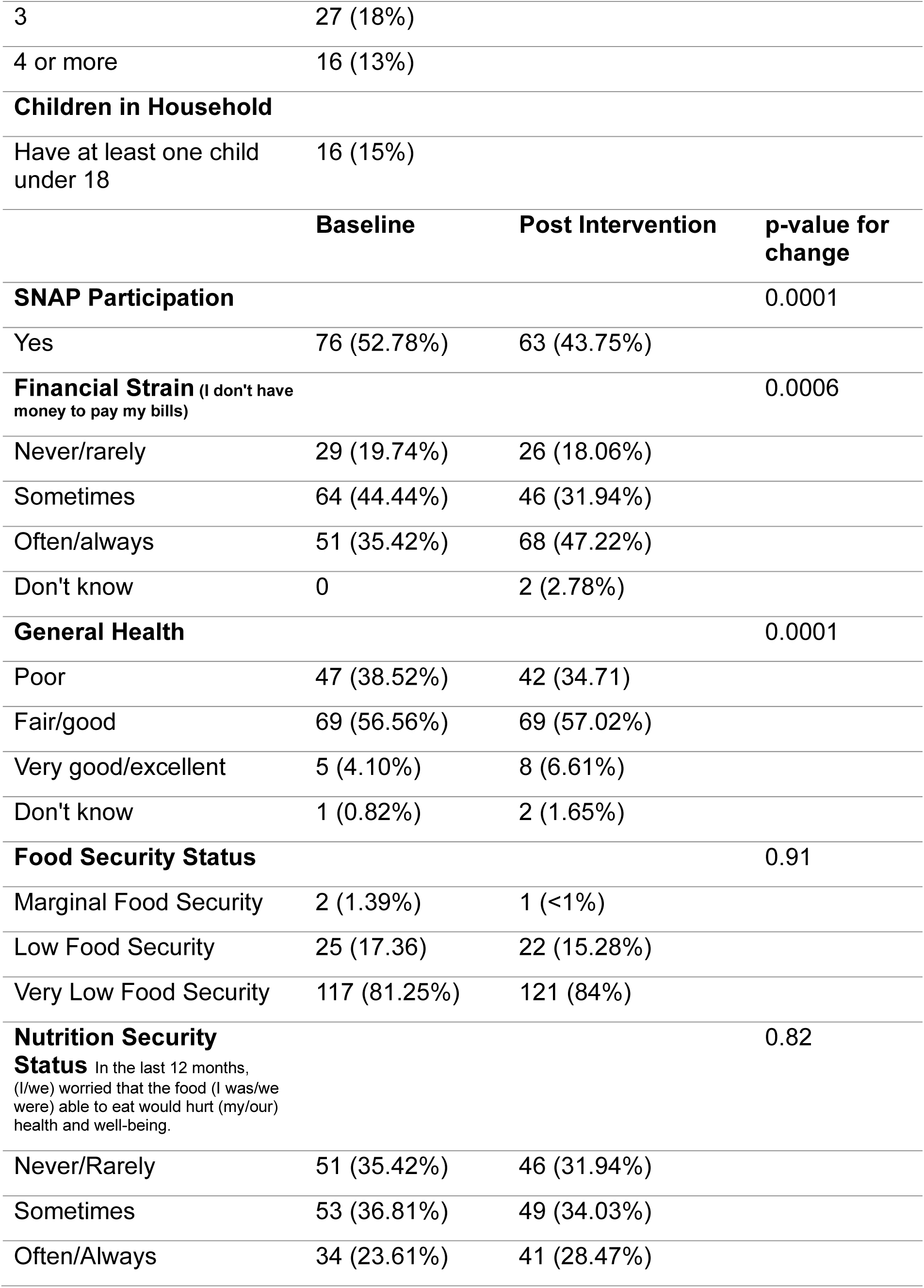
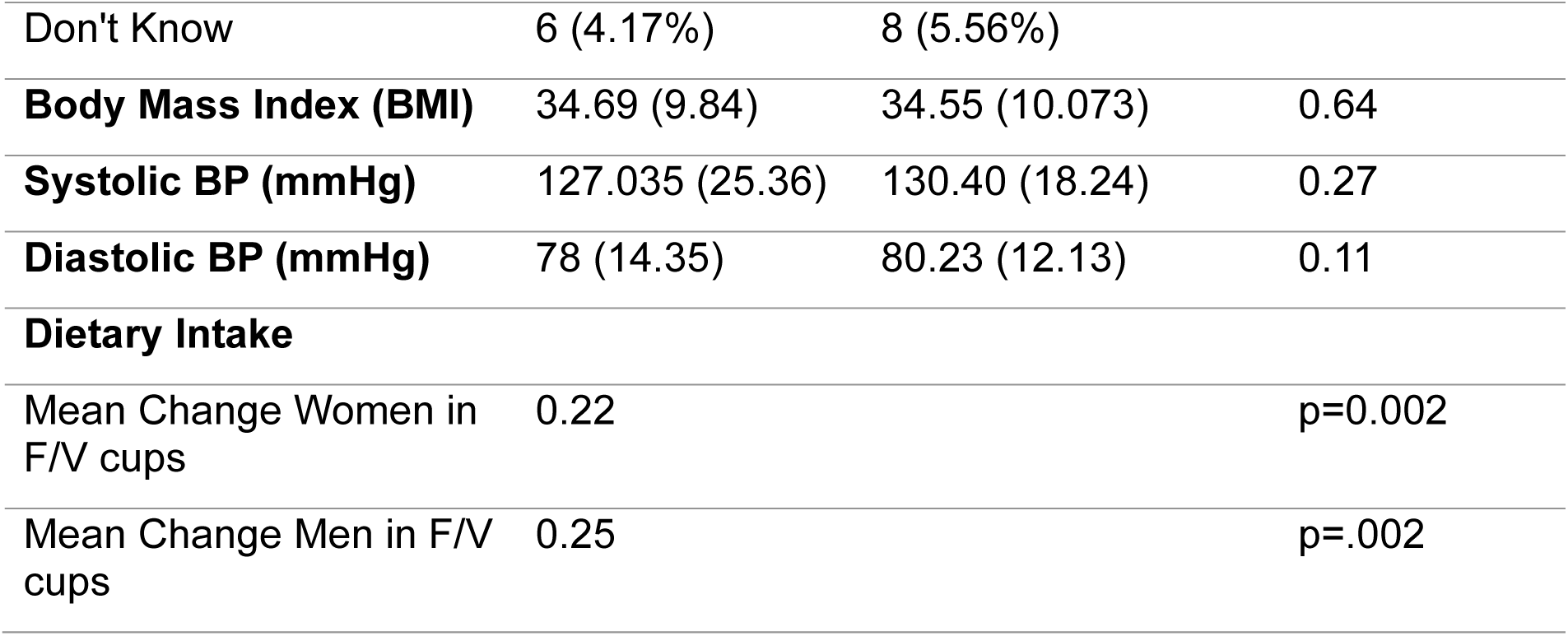
Demographics of Study Participants and Pre/Post Results Across the Intervention Period.

|  | **Baseline**<br>**(N=144) N (%)** |
| --- | --- |
| **Sex** |  |
| Female | 90 (62.50%) |
| **Age (years)** | 52.97 (SD=9.00) |
| **Race/Ethnicity** |  |
| White | 119 (82.64%) |
| Black or African American | 22 (15.28%) |
| Hispanic or Latino | 2 (1.39%) |
| **Education Level** |  |
| Less than HS | 42 (29.17%) |
| HS Graduate/GED | 78 (54.17%) |
| College Graduate | 24 (16.67%) |
| **Employment Status** |  |
| Employed | 32 (22.22%) |
| Not Employed/Other | 112 (77.78%) |
| **Household Income** |  |
| Low Income (<\$25k) | 103 (71.53%) |
| Middle Income (\$25k-\$74k) | 31 (21.53%) |
| High Income (≥\$75k) | 5 (3.47%) |
| Prefer not to answer | 5 (3.47%) |
| **Household Size** |  |
| 1 | 44 (30%) |
| 2 | 57 (39%) |

|  |  |  |  |
| --- | --- | --- | --- |
| 3 | 27 (18%) |  |  |
| 4 or more | 16 (13%) |  |  |
| **Children in Household** |  |  |  |
| Have at least one child under 18 | 16 (15%) |  |  |
|  | **Baseline** | **Post Intervention** | **p-value for change** |
| **SNAP Participation** |  |  | 0.0001 |
| Yes | 76 (52.78%) | 63 (43.75%) |  |
| **Financial Strain** (I don't have money to pay my bills) |  |  | 0.0006 |
| Never/rarely | 29 (19.74%) | 26 (18.06%) |  |
| Sometimes | 64 (44.44%) | 46 (31.94%) |  |
| Often/always | 51 (35.42%) | 68 (47.22%) |  |
| Don't know | 0 | 2 (2.78%) |  |
| **General Health** |  |  | 0.0001 |
| Poor | 47 (38.52%) | 42 (34.71%) |  |
| Fair/good | 69 (56.56%) | 69 (57.02%) |  |
| Very good/excellent | 5 (4.10%) | 8 (6.61%) |  |
| Don't know | 1 (0.82%) | 2 (1.65%) |  |
| **Food Security Status** |  |  | 0.91 |
| Marginal Food Security | 2 (1.39%) | 1 (<1%) |  |
| Low Food Security | 25 (17.36) | 22 (15.28%) |  |
| Very Low Food Security | 117 (81.25%) | 121 (84%) |  |
| **Nutrition Security Status** In the last 12 months, (I/we) worried that the food (I was/we were) able to eat would hurt (my/our) health and well-being. |  |  | 0.82 |
| Never/Rarely | 51 (35.42%) | 46 (31.94%) |  |
| Sometimes | 53 (36.81%) | 49 (34.03%) |  |
| Often/Always | 34 (23.61%) | 41 (28.47%) |  |
| Don't Know | 6 (4.17%) | 8 (5.56%) |  |
| **Body Mass Index (BMI)** | 34.69 (9.84) | 34.55 (10.073) | 0.64 |
| **Systolic BP (mmHg)** | 127.035 (25.36) | 130.40 (18.24) | 0.27 |
| **Diastolic BP (mmHg)** | 78 (14.35) | 80.23 (12.13) | 0.11 |
| **Dietary Intake** |  |  |  |
| Mean Change Women in F/V cups | 0.22 |  | p=0.002 |
| Mean Change Men in F/V cups | 0.25 |  | p=.002 |

**Table 4.** Primary and Secondary Outcomes Changes within MTM and Grocery Rx Program and across Supplemental Nutrition Assistance Program (SNAP)

|  | **MTM (n=33)** | **Grocery Rx (n=111)** |
| --- | --- | --- |
|  | ***M Change (CI)*** | ***M Change (CI)*** |
| **Systolic (mmHg)** | 9.39 (-3.99, 22.78) | .25 (-3.57, 4.08) |
| **Diastolic (mmHg)** | 5.45 (-3.61, 14.51) | 1.26 (-1.16, 3.69) |
| **Dietary Intake (Fruits and vegetable cups)** |  |  |
| Men | .49 (.20, .77)* | .24 (.08, .40)* |
| Women | .24 (.04, .45)* | .22 (.11, .33)* |
| **General Health** | 7.71 | 48.65* |
| **Financial Strain** | 9.08 | 17.83* |
|  | **MTM + SNAP (n=23)** | **Grocery Rx + SNAP (n=53)** |
| **Systolic (mmHg)** | -13.77 (-58.15, 30.59) | -8.75 (-16.83, -.67)* |
| **Diastolic (mmHg)** | -9.30 (-39.23, 20.62) | -5.42 (-10.72, -.13)* |
| **\*Statistically significant result at $p < .05$** |  |  |

Between baseline and post-intervention across all groups, there was a significant change in SNAP participation, financial strain, and general health status.

Table 4. indicates that among the MTM or grocery Rx participants there was not a significant change in systolic or diastolic blood pressure. However, across the secondary outcomes there was a significant increase in fruit and vegetable cups for both men and women across both groups. Within MTM, both men and women increased their intake by about 1/5^th^ of a cup. In Grocery Rx, men increased by ½ cup and women increased by ¼ cup. There was also a significant change in general health and a decrease in financial strain among those in the grocery Rx program. Results also indicate that those who participated in SNAP at the beginning of the program had a significant decrease in systolic and diastolic blood pressure among the grocery Rx participants compared to those not participating in SNAP.

### Cost Efficiency Results

The intervention specific cost differences are primarily driven by the cost of food and postage. Participants in the MTM arm of the study received bi-weekly deliveries at a cost of $181 per month inclusive of shipping costs. In the Grocery Rx (urban) arm, participants received a card directly from Soda Health which was allocated $100 dollars each month of the study. The Grocery Rx (rural) arm utilized physical vouchers, also in the amount of $100, which were mailed to participants once per month, incurring additional postage costs and additional staff time to prepare the packages. The Grocery Rx (online) arm utilized online shopping with account credits for $100 each month; in addition to the amount available to purchase groceries, the Instacart arm was provided an additional $10 to cover service fees. While MTM participants received the full value of the meals provided, grocery Rx participants utilized, on average, 84.1% of the funds allocated. Cost calculations include the full allocation provided to participants to avoid understating the potential costs of implementation.

As may be expected, costs of the 3 grocery Rx interventions are similar at less than $400 per participant. There were small differences attributed to the additional funds to cover fees from Instacart (online arm), the additional shipping costs associated with sending vouchers for Food City (rural arm), and the lack of shipping costs covered by Soda Health (urban arm) to send shopping cards to participants. For all the grocery Rx programs, more than 75% of the total costs were available to participants to spend on food, a percentage that would grow as the intervention was extended. The MTM arm of the intervention cost more than the grocery Rx arms at just over $600 for the 3-month period for each participant. The difference between home delivery of 20 frozen meals per month and access to $100 in groceries makes direct comparisons difficult. However, the value of the food and delivery ($543 for 3 months) accounted for 90% of the total costs of that arm of the study.

### Semi-structured Qualitative Interview Results

A total of 22 participants were contacted about participating in interviews, and 20 interviews were completed (91% response rate). The participant sample sizes from each intervention group included: six MTM, six grocery Rx (rural), and eight grocery Rx (urban). Four main themes were identified from the interview data, including: user satisfaction, user recommendations, communication and nutrition education, and health and behavioral outcomes. The main themes were consistent across participants from each of the three groups (MTM, grocery Rx rural, and grocery Rx urban), so the results are presented collectively with relevant divergence among groups noted in Table 5.

**Table 5.**
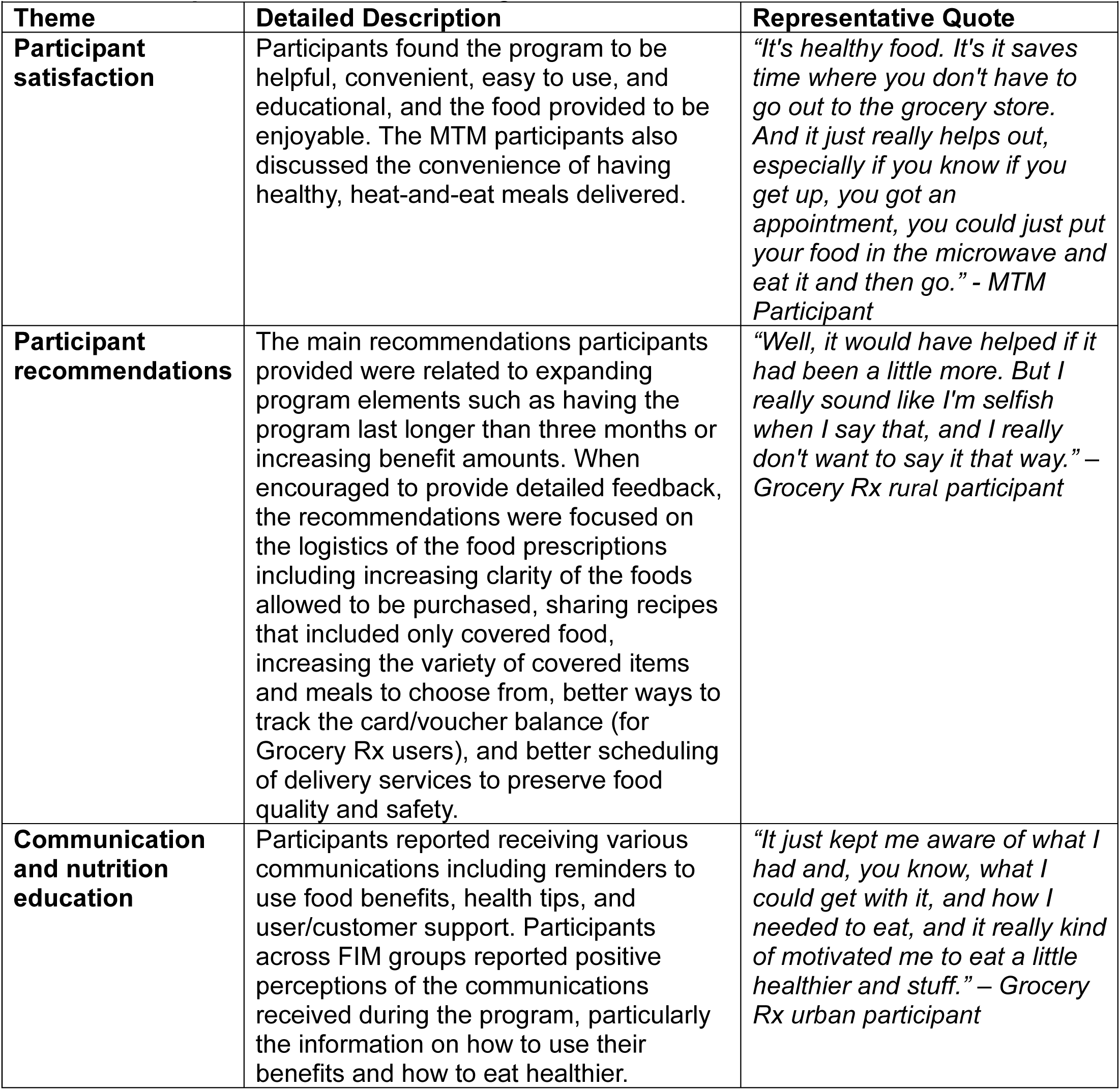
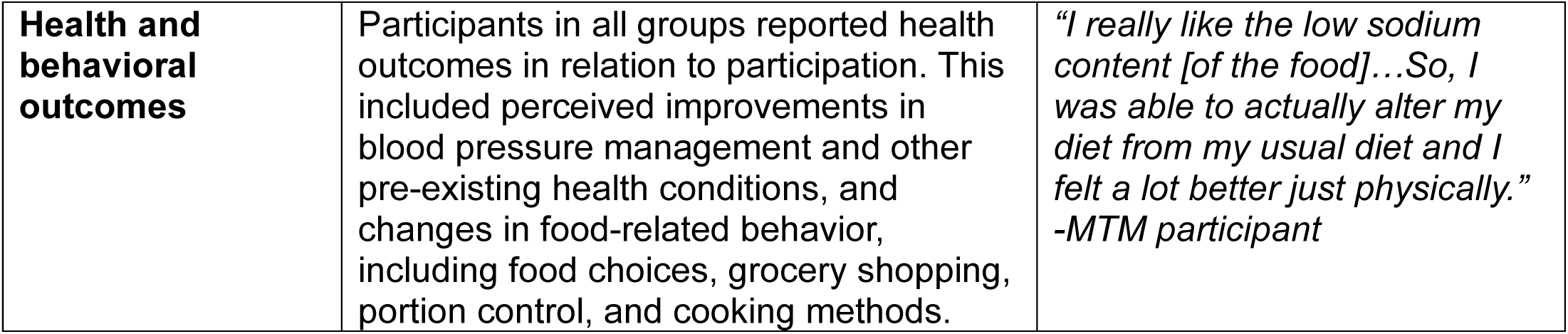
Participant Semi-structured Program Feedback Interview Themes.

Participants reported being motivated to participate in the program for the potential health benefits, due to being food insecure/needing food assistance, and because the program was easy to participate in. Table 5. contains detailed descriptions of each theme and representative quotes.

## Discussion

Our study is impactful in two meaningful ways from a process measures and implementation standpoint. First, our participants redeemed 84% of their grocery benefits and 99.5% of intended meals were delivered to participants in the MTM. Relative to other studies, this is a high redemption rate and completion rate across our lower income rural and urban adults. Other studies have reported a redemption rate ranging from below 20% to 80% or greater,^33^with one study indicating an overall voucher redemption rate of 52%^22^. However, our results are similar to other grocery Rx programs, such as the Fresh Rx produce grocery program, which had a rate of 79%^23^. The average adult participant spent 73.1% of their produce prescription dollars during program enrollment (the remainder was unspent)^34^. A similar FIM program in rural and urban areas of Georgia had a 59% retention rate at 6-months,^35^ compared to our engagement and retention rates of 95% and 91%, respectively. We attribute the retention and engagement rate to utilizing a screening decision tool to tailor the intervention to participants rather than conducting a conventional randomized controlled trial. In addition, we had dedicated staff to provide high touch patient care and a text help line.

Over 12 weeks in both MTM and grocery Rx programs, there was no significant change in blood pressure. However, our study did not have a comparison group and thus we cannot make any comparisons between our study relative to standard care. Moreover, our study was not powered to detect changes in blood pressure but was designed to seek information related to tailoring a FIM program for user needs, preferences, and geographic location to retain engagement. In our qualitative interviews, participants reported improved symptoms related to cardiovascular disease including reduced swelling, feeling better overall, and being able to manage their blood pressure better.

Previous studies have found mixed results of effects of FIM on blood pressure, with some studies finding a reduction in systolic and diastolic blood pressure^34^ among adults with HTN utilizing a produce prescription program or a grocery voucher program,^15^ while another yielded no change in blood pressure between treatment and control over a 6-month program^36^. Our results, coupled with these previous findings, point to the need for larger scale studies utilizing a user-centered approach to allocate individuals based on their level of need, stage of disease, and preferences in order to understand for whom, how long, and in what manner a FIM program can improve clinical outcomes.

Our secondary outcomes indicated strong improvements in dietary intake, general health, and financial strain. While this is a small change, it is also clinically meaningful and points to how a tailored dietary program can “move the needle” on key health outcomes. Other FIM programs have reported similar findings with general health and dietary intake^34^. Our study further expands this research to suggest that a short-term intensive FIM program designed for the user can improve diet and feelings of improved health in the short term. Participants self-reported improved dietary habits such as eating more fruits and vegetables, eating foods that are beneficial for managing chronic conditions like diabetes, and being able to purchase healthful foods or meals they would not be able to purchase without the benefits they were provided.

Our study also explored how SNAP participation, in addition to receiving a tailored FIM program, influenced blood pressure change. Our results point to the effects that dual participation in SNAP and a grocery Rx program can have on reducing blood pressure. While this result was not powered, the study shines a light on the role that additional supplemental funds can have for certain groups. Studies to date have indicated that SNAP has been associated with unhealthy purchases,^37,38^ while others have indicated that participation reduces food insecurity and lowers annual health care expenditures^39^. Our preliminary findings corroborate previous findings suggesting that, in the short-term, SNAP + FIM can ease financial strain, reduce stress, and free up cognitive bandwidth, which all in turn allows individuals to make healthier choices ^39^. A recent scoping review indicated that nutrition education + monetary incentives among SNAP participants resulted in the greatest improvements for dietary intake^40^. Our results further support the notion of offering SNAP participants an additional supplemental program for purchasing healthy food that meets their individual needs. This can allow participants an opportunity to free up financial resources while also engaging in a program that allows for learning how best to utilize their SNAP benefit in a healthful manner^41^.

Our study had several limitations worth noting. Participation was only for 3 months, limiting the ability to assess long-term effects. In addition, there was no tailoring based on the level of food security and disease management. This is a critical piece in helping to inform who and at what level these clinical-community linked resources are most appropriate. For some participants, the health metrics provided by clinics did not coincide with the end of the program (3 months post-baseline). As a result, health measures for post-intervention were self-reported by 17 participants. Moreover, qualitative interviews were only conducted with program completers who volunteered to do an extra interview and thus may have had a more favorable perception of the program than others who did not complete the program or did not want to participate in additional data collection.

## Non-standard Abbreviations and Acronyms

AHA: American Heart Association
EMR: Electronic medical record
FIM: Food is Medicine
GusNIP: Gus Schumacher Nutrition Incentive Program
HCxF: Health Care by Food
MTM: Medically tailored meals
PHI: Protected health information
SNAP: Supplemental Nutrition Assistance Program

## Data Availability

Data can be available once publication has been accepted Dataset DOI: 10.5061/dryad.xsj3tx9ss Description of the data and file structure

## Acknowledgements

The study team would like to acknowledge all the study participants in providing their time and support in our user-centered design food is medicine intervention. We would also like to acknowledge all the clinic staff that were involved in screening and referral of patients into the Healthcare by Food system.

## Supplemental Materials

Screening Decision Tool

